# Faecal immunochemical testing (FIT): Sources of analytical variation based on three years of routine testing in the context of DG30

**DOI:** 10.1101/2020.04.15.20066191

**Authors:** Tim James, Brian D Nicholson, Rhiannon Marr, Maria Paddon, James E East, Steve Justice, Jason L Oke, Brian Shine

## Abstract

**Aims:** To determine analytical capabilities of a commonly used faecal immunochemical test (FIT) to detect haemoglobin (Hb) in the context of NICE guidance DG30, and the likely use of FIT to reprioritise patients delayed by the COVID-19 pandemic.

**Methods:** Data obtained from independent verification studies and clinical testing of the HM-JACKarc FIT method in routine primary care practice were analysed to derive analytical performance characteristics.

**Results:** Detection capabilities for the FIT method were 0.5 µg/g (limit of blank), 1.1 (limit of detection) and 15.0 µg/g (limit of quantification). 31 of 33 (94%) non-homogenised specimens analysed in triplicate were consistently categorised relative to 10 µg/g compared to all 33 (100%) homogenised specimens. Imprecision in non-homogenised specimens was higher (median 27.8%, (range 20.5% - 48.6%)) than in homogenised specimens (10.2%, (7.0 to 13.5%)). Considerable variation was observed in sequential clinical specimens from individual patients but no positive or negative trend in specimen degradation was observed (p=0.26).

**Conclusions:** The FIT method is capable of detecting Hb at concentrations well below the DG30 threshold of 10 µg/g. However, total imprecision is considerable when including sampling variation. Binary categorisation against a single defined threshold above and below 10 µg/g was more consistent and improved following specimen homogenisation. This approach may be more appropriate when reporting results for symptomatic patients tested in primary care, including those who have had definitive investigation delayed by the COVID-19 pandemic and need to be re-prioritised.

**Key Messages:** Faecal immunochemical testing (FIT) is increasingly used to detect blood at low haemoglobin (Hb) concentrations in specimens from symptomatic primary care patients but the analytical characteristics in this context have not been fully documented.

A commonly used FIT method showed good capability in a routine UK clinical setting to detect Hb at the NICE recommended threshold of 10µg/g. Imprecision estimates were considerable when sampling was considered, even above the limit of quantification of 15 µg/g.

Analytical variability appears too high for reliable reporting of quantitative Hb concentrations: reporting positive or negative results around a threshold of 10µg/g appears more appropriate after sample homogenisation.

Dichotomous FIT reporting is likely to be an important tool to risk stratify patients with lower GI cancer symptoms who have had their test deferred due to the COVID-19 pandemic

## Introduction

Colorectal cancer is globally the third most incident malignancy (1). It is surgically treatable with improved long-term outcomes if diagnosis is at an early stage (2). Most developed countries, including the UK, operate colorectal screening programmes using faecal occult blood testing. Screen detected cancers benefit from early diagnosis and treatment, with associated improved survival (2). Faecal immunochemical tests (FIT) have largely replaced the traditional guaiac based faecal occult blood tests due to the increased specificity of FIT.

To complement the UK Bowel Cancer Screening Programme (UKBCSP), the 2017 DG30 NICE guidance (3) recommended the use of FIT for faecal haemoglobin (Hb) detection in patients presenting to primary care with low risk abdominal symptoms. The adoption of FIT in primary care has been slow, with notable variation in uptake and implementation across the UK (4). The Oxford University Hospitals Trust (OUH) adopted FIT prior to the DG30 guidance to comply with the 2015 NG12 NICE guidance for suspected cancer (6), which recommended the use of faecal occult blood testing in symptomatic patients. This coincided with a desire from the clinical laboratory to move away from the comparatively inaccurate guaiac based method. FIT was commissioned by Oxfordshire Clinical Commissioning Group (OCCG) as a direct access test for General Practitioners in 2016 following a study by this group comparing the accuracy of the guaiac and FIT methods in symptomatic primary care patients meeting NG12 criteria (5).

Despite gradual uptake of FIT, there remains a clear need to understand FIT method characteristics (7). FIT testing, whether undertaken within a screening programme or applied to a symptomatic population, is dependent on the analytical performance of the laboratory procedures used. These analytical characteristics, most crucially sampling, impact on the way the results are most accurately reported (8). Whilst there has been significant work undertaken in the context of screening (9) there is less data available in the symptomatic population. This is important to explore as the characteristics of the population for a screening programme, and the associated specimens, may differ from patients presenting with symptoms. For example, the age-range of the symptomatic population is broader and specimen characteristics, including faecal consistency, will likely differ in patients with changes in bowel habit, and the information and support for taking a sample is less. This may affect sample, and sampling integrity.

The FIT methods used to process samples from symptomatic patients are those developed for measuring faecal Hb at higher concentrations in screening. At present, the cut-off used by the UKBCSP is >120 µg/g, with future plans to reduce this to approximately 50 µg/g to improve sensitivity. The threshold recommended for use with symptomatic patients in DG30 is 10 µg/g. FIT methods may not be optimised, or fully characterised, for detection at the lower concentrations now being considered important for application in DG30 (7).

While introduction of FIT into primary care cancer pathways is limited at present, given the large backlog of endoscopy created by the COVID-19 pandemic it is almost certain the widespread use of low cut off FIT testing will be introduced immediately the pandemic has passed to risk stratify all patient referred with possible cancer symptoms into groups for urgent and less urgent endoscopy. Understanding low cut off FIT test performance in the context of symptomatic patients is therefore a priority.

In the present study we present our observations and assessment of FIT method performance since introduction of the test into routine clinical practice for a symptomatic primary care population over a three year period. We include estimates of analytical performance, observations on longer term method performance, sampling reproducibility in homogenised compared to non-homogenised material, and consistency is sequential specimens from the same patients.

## Methods

### Setting and analytical method

A single laboratory undertaking centralised analysis of FIT requests, mainly from primary care, for the 680,000 population of Oxfordshire.

FIT analysis was undertaken using the HM-JACKarc analyser (Kyowa Medex Co., Ltd., Tokyo, Japan) (ref Guildford report) which has a calibration range of 7 to 450 µg Hb/g faeces. This method was introduced into service prior to NICE guidance defining 10 µg/g as the threshold defined for detection and we used 7 µg/g, the lowest calibrator value as the threshold for a positive result

All specimens were requested by NHS primary care clinicians and collected into standard stool pots by patients, a sampling approach taken due to concerns about sampling capability when undertaken by (often elderly) patients (5). Clinician advice from OUH and OCCG included guidance on delivering the sample to the laboratory on the same day as collection as Hb can degrade on storage (10). On arrival in the laboratory the stool specimens were sampled using the Extel Hemo-Auto MC designed for application with the HM-JACKarc by laboratory staff competency assessed in this technique. The whole sampling procedure was undertaken in a fume cupboard by staff with standard laboratory protective equipment and involved a 30 second vortex mix which for most specimens was adequate for release of the faecal pellet collected into the dimples/grooves of the picker. All specimens were visually inspected to ensure complete suspension of specimen and if it was noted any residual specimen adherence mixing continued until all material was removed.

### Method detection capability estimates and immunoassay reproducibility

The FIT method limit of blank (LOB), limit of detection (LOD) and limit of quantification (LOQ) were estimated as recently recommended (7). As it was not possible to obtain a genuine faecal sample, assured as having no Hb analyte present, sample blank readings were utilised to estimate LOB. LOD was calculated using the LOB + 1.645 (SD) of samples with “very low concentrations”, considered to be those giving FIT results those between 0.5µg/g an 1.0 µg/g.

LOQ was estimated from an imprecision profile constructed from faecal samples analysed in duplicate across a wide FIT concentration range of naturally Hb positive material and was taken as the concentration at which the percent coefficient of variation (%CV) was 10%. A 10%CV was selected as this has been recommended as an appropriate imprecision threshold for FIT method LOQ evaluation (7). An additional within batch imprecision estimate was made using a dilution of the supplier’s internal quality control (IQC) material. This was prepared to give a concentration near to the anticipated LOD to assess whether the behaviour of IQC material in the assay system with respect to imprecision was similar to true samples.

The on-instrument intermediate imprecision (*not* including sampling into the collection device) was estimated from two level IQC materials provided by the manufacturer over several months to include the effects of different analysts, reagent lot numbers and calibration material lot numbers.

### Effects of sample homogenisation

It is unclear whether Hb is evenly distributed in faeces therefore we investigated the Hb concentration before and after sample homogenization. Randomly selected faecal specimens arriving in standard stool containers were sampled in triplicate, homogenized and then resampled a further three times. This resulted in six estimates of the result (3 non-homogenised replicates and 3 homogenised replicates) with the imprecision presented as the %CV of each set of three replicates. Individual stool specimen replicates were sampled by the same member of staff but overall three different members of laboratory staff were involved in this part of the study.

### Stability of specimens within the collection device

FIT analysis was undertaken as a batch once or twice per week. To verify the stability of Hb in the buffered collection device a selection of specimens were analysed at three time points (at time zero, at 6 days and at 21 days).

### Within patient serial sampling

Prior to January 2016 the standard practice with the guaiac method was to collect serial samples. Despite revised guidance, recommending only a single specimen was required for FIT, many primary care clinicians continued to request more than one. This may have been due to clinicians being unaware of revised guidance or because there remained doubt about test credibility and the additional reassurance multiple specimens might provide (11,12). Sequential samples were delivered to the lab on the same day allowing assessment of delayed analysis on FIT concentration. These sequential specimens represented maximum within patient error as Hb concentrations reflected total variability (between day biological variation, analyte instability, sampling error and immunoassay method imprecision).

### Statistical analysis

Differences between the imprecision of sampling between non-homogenised and homogenised sampling were assessed using a Wilcoxon rank test with continuity correction, the null hypothesis being that the distribution of × – y (%cv in homogenised group – %cv in non-homogenised group) is symmetric about 0.

For patients with multiple specimens and discordant results (that is, at least one result greater than, and at least one result less than 10 ug/g), we examined whether there was an association between the likelihood of a positive result and delay in processing the specimen in the laboratory of 48 or 72 hours. In this group, we also examined whether there was an association between a positive result and the sequence number of the specimen. Both were assessed using the Chi-squared test.

## Results

### Detection capabilities and immunoassay reproducibility

#### Limit of the blank (LOB)

The mean FIT result of the sample blank run as part of the method IQC over 100 consecutive batches over 11 months and three different reagent batches was 0.20 µg/g, SD 0.16 µg/g, from which a derived LOB was 0.47 µg/g.

#### Limit of detection (LOD)

As the method only reports to one decimal place we considered this to be 0.5 µg/g for practical purposes. 51 samples with apparent concentrations between 0.5 and 1.0 µg/g measured in duplicate were considered “very low concentrations” and suitable for calculation of an SD for the LOD calculation. The derived SD was 0.35 µg/g from which an LOD of 1.1 µg/g was derived (from LOB + 1.645SD).

#### Limit of quantification (LOQ)

LOQ was calculated from a precision profile (Figure 1) constructed using data from 132 paired, within-batch replicates, analysed in seven different batches over a one week period. Three samples with concentrations above the highest calibrant concentration, 450 µg/g, were excluded from the analysis. The imprecision was 13% at approximately 7 µg/g (the lowest calibrator value), 11.5% at 10 µg/g (The NICE threshold for detection) and 10% at 15 µg/g. The 10% imprecision threshold has been recommended as the LOQ for FIT (7) and therefore 15 µg/g was considered to be this assay characteristic.

**Figure 1 legend:**
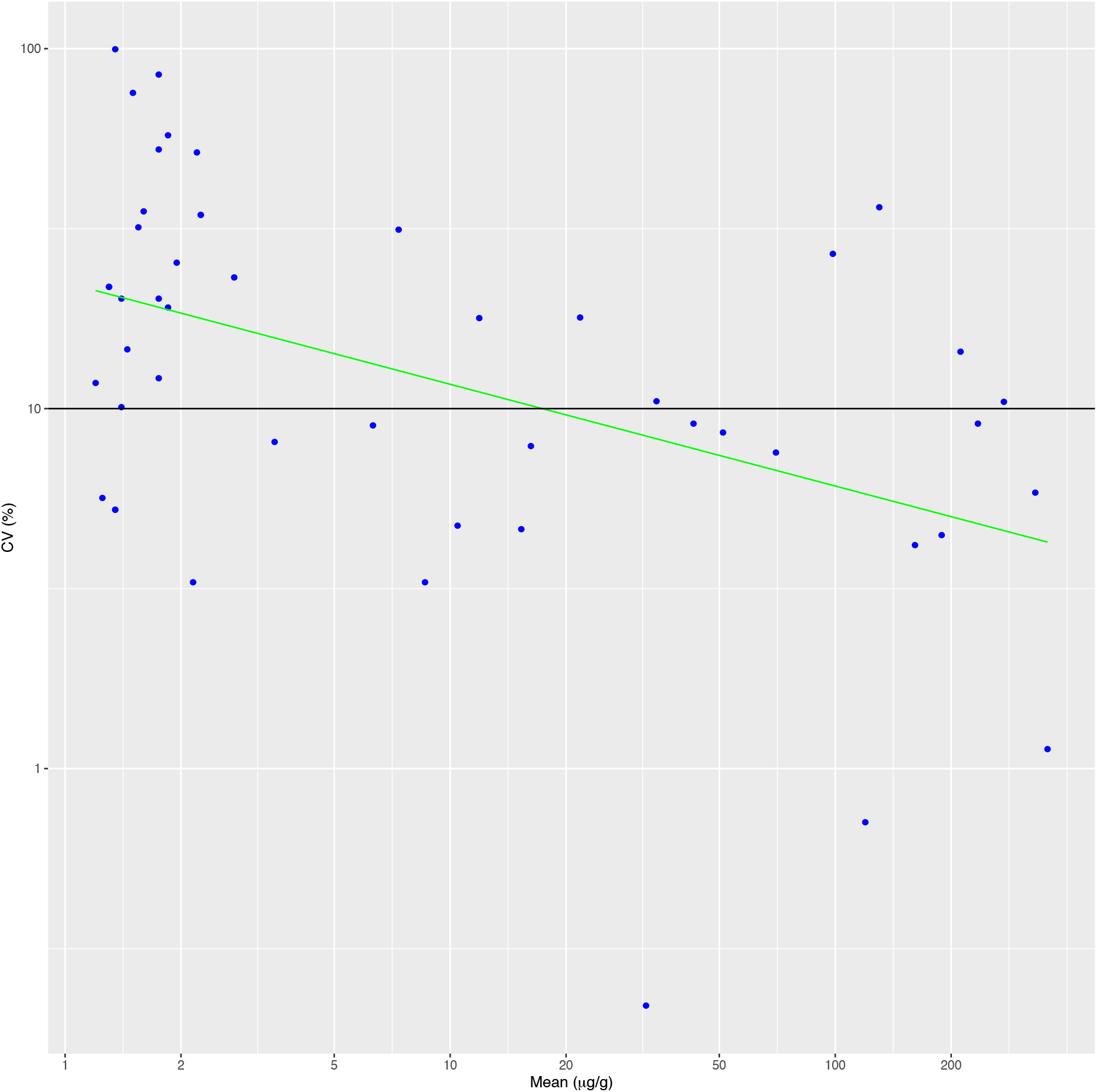
Imprecision profile for FIT analysis. The 10% CV limit used to define the limit of quantification (LOQ) is shown.

#### Method precision and reproducibility

The within-batch imprecision characteristics of a highly diluted IQC material were mean 1.5 µg/g, SD 0.5 µg/g, %CV 33.9 %. Estimation of on-instrument method precision utilised quality control data collected over 12 months during which time four different lot numbers were used (table 1).

**Table 1:**
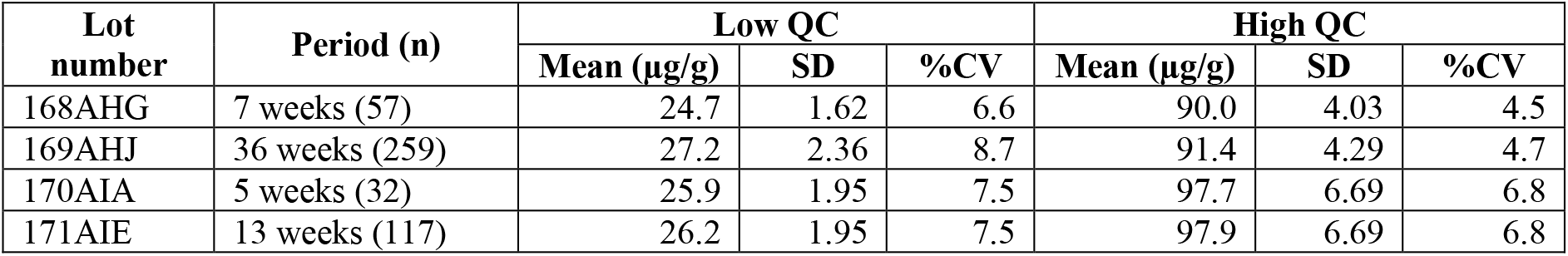
Imprecision estimates over a 12 month period

| Lot number | Period (n) | Low QC |  |  | High QC |  |  |
| --- | --- | --- | --- | --- | --- | --- | --- |
|  |  | Mean (µg/g) | SD | %CV | Mean (µg/g) | SD | %CV |
| 168AHG | 7 weeks (57) | 24.7 | 1.62 | 6.6 | 90.0 | 4.03 | 4.5 |
| 169AHJ | 36 weeks (259) | 27.2 | 2.36 | 8.7 | 91.4 | 4.29 | 4.7 |
| 170AIA | 5 weeks (32) | 25.9 | 1.95 | 7.5 | 97.7 | 6.69 | 6.8 |
| 171AIE | 13 weeks (117) | 26.2 | 1.95 | 7.5 | 97.9 | 6.69 | 6.8 |

Method reproducibility, expressed as %CV, for the low IQC material with a concentration of around 26 µg/g ranged from 6.6% to 8.7% and for the high IQC material, concentration around 90 µg/g ranged from 4.5% to 6.8%.

### Effects of sample homogenisation

31 (94%) of 33 non-homogenised samples were consistent with respect to their categorisation as above or below the 10 µg/g (figure 2). 27 had a mean result below 10 µg/g (negative) and 6 had mean results above (positive). Four had all three replicates above the LOQ and imprecision estimates were calculated: median %CV of 27.8%, range 20.5% to 48.6%.

**Figure 2 legend:**
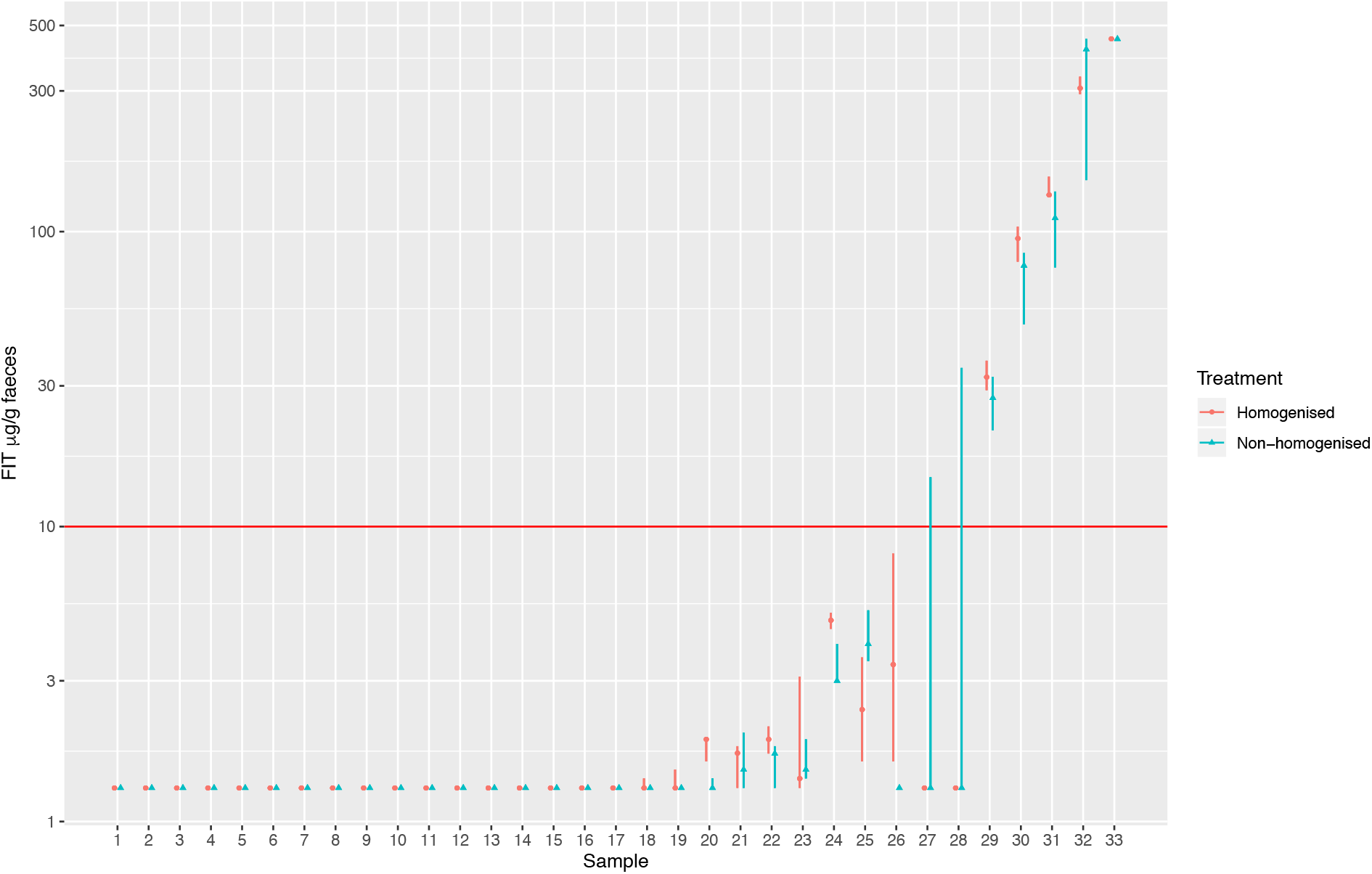
Specimen preparation effects. 33 specimens analysed in triplicate prior to and after homogenisation. Results show the observed concentration range (lowest to highest) with each specimen presented sequentially with homogenised (circles) and non-homogenised (triangles) results adjacent.

Two further samples had two replicates below the LOQ which prevented accurate calculation of imprecision but had discrepant result relative to the 10 µg/g NICE threshold. One specimen had replicates of <1.1, <1.1, 34.5 µg/g (two below the LOD and one above the LOQ). Clinical details of this patient were PR bleeding and endoscopy showed an 8mm polyp in the sigmoid. Histology revealed the polyp to be non-dysplastic and hyper-plastic and this patient was discharged. The second of these discrepant results had replicates <1.1, <1.1, 14.7 µg/g (two below the LOD and one between the LOD and LOQ and positive relative to the NICE threshold of 10 µg/g). This patient was being followed up by an established vague symptoms pathway (13) and whilst nothing abnormal was detected after 9 months of follow-up, at the time of patient assessment they were noted to be taking NSAID.

In the homogenised specimen group, all 33 (100%) specimens were consistent with respect to their categorisation as above or below the 10 µg/g (figure 2). 28 had a mean result below 10 µg/g (negative) and 5 had mean results above (positive). Categorisation as positive or negative results was consistent in all replicates relative to a threshold of 10 µg/g. Four of the specimens in the homogenised set had all three replicates above the LOQ and imprecision estimates were calculated: median %CV of 10.2%, range 7.0 to 13.5%. This was lower but not statistically significant (*p = 0*.*10*) than the imprecision observed in the non-homogenised samples.

### Stability of specimens within the collection device

15 specimens selected as their initial result was above the LOD, (8 below the LOQ and 7 above) were repeat tested. All 15 specimens had consistent categorisation when repeat tested at 6 days compared to the initial result (positive or negative relative to the 10 µg/g threshold). At 21 days all but one specimen were consistently categorised however, however one had fallen from 25 to 6 µg/g. For the 7 specimens above the LOQ there was no statistical difference at 6 (p=0.95) or 21 days (p=0.93).

#### Within patient serial sampling

524 patients sent more than one sample within a 3 month period: 302 returned 2 specimens; 222 returned 3; 2 returned 4; and 1 returned 5. Of the 225 patients with 3 or more specimens, 197 (87.6%) had concordant results for all specimens (188 negative, 9 positive). 28 (12.4%) had discordant results, that is, at least one value more, and at least one value less, than 10 µg/g.

There was no obvious trend on visual review of plots of individual patient’s FIT values over time (figure 3), and no association between the likelihood of a positive result based on either a delay of more than 2 days in the specimen reaching the laboratory (p = 0.84, Chi-squared test). There was no association between the likelihood of a positive results and the number in the sequence of a particular patient’s specimens (p = 0.21, Chi-squared test),

**Figure 3 legend:**
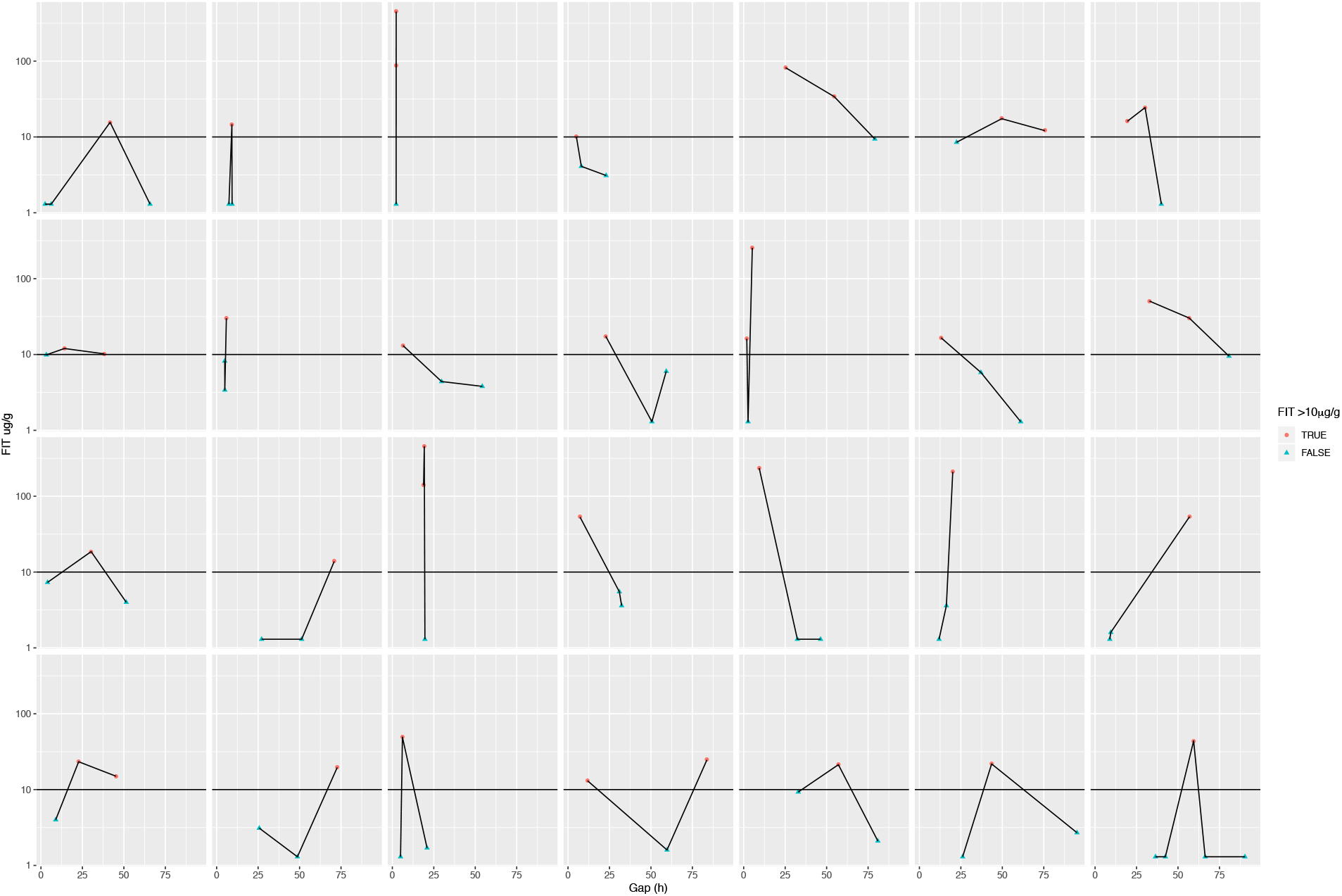
Within patient variation of results in the 28 patients whose serial samples were discordant with respect to categorisation as positive (>10 µg/g) or negative (<10 µg/g). Each panel represents a single patient, time (hours) vs concentration (µg/g) profile showing the inconsistent patterns observed.

## Discussion

Current proposals for FIT testing at low concentrations are dependent on accurate LOD and LOQ estimates (7). In this evaluation we have derived these detection characteristics for a commonly used FIT method. The LOB (0.5 µg/g), LOD (1.1 µg/g) are considerably lower than the Hb concentration of 10 µg/g designated by NICE as the threshold for further patient investigation. The LOQ, if defined as the Hb concentration where imprecision is 10%CV (7) was observed to be 15.0 µg/g. These detection capability estimates would suggest that at 10 µg/g the method is sensitive enough to be confident that Hb is present but not about the absolute value below 15 µg/g.

The immunoassay method imprecision is good when assessed using non-clinical IQC material, being consistently <9%, even over an 8 month period. A diluted IQC material at a concentration (1.5 µg/g) near to the LOD (1.1 µg/g) showed an imprecision of 33.9%, higher than the precision profile line of fit derived with clinical material but reflective of highly variable imprecision at these lower concentrations. The concentration of Hb present in the two IQC materials provided by the manufacturer, at around 25 µg/g and 95 µg/g. These were presumably selected to be clinically appropriate for use in screening programmes where decisions thresholds are higher. When the clinical interest shifts to lower concentrations, we would suggest a strategy of using IQC materials with values below and above 10 µg/g.

Imprecision rises when faecal sampling is taken into consideration: average imprecision in homogenised specimens was 10.2% and in non-homogenised specimens was 27.8%, with upper estimates of 13.5% and 48.6%, respectively. Additionally, inconsistent classification was observed with respect to the 10 µg/g threshold in two further samples not assessed for imprecision due to some or all of the values being below the LOQ. This would suggest that Hb is not uniformly distributed in the faecal specimen, a finding consistent with other faecal constituents. For example, a ^1^H NMR spectroscopy study of faeces (14) highlighted both heterogeneity and instability of a range of metabolic constituents. Also, whilst there is limited evidence to support the practice it is common for specimens for faecal elastase and calprotectin to include a homogenisation step prior to analysis. Future work must further establish the potential benefit of homogenisation in relation to FIT. We note that, in two specimens, one of the three non-homogenised results was positive compared with none in the homogenised specimens. This might lead to speculation that homogenisation leads to false negative results by reducing the amount of Hb to the average in the stool specimen. Neither of these patients have had a diagnosis of colorectal cancer, more than 1 year after the specimens were first obtained.

The stability of the Hb in the sampling device buffer appears reasonable at 6 days and for most specimens up to 21 days. However, one specimen showed notable deterioration at 21 days from above to below the current 10 µg/g threshold. In a DG30 service setting a minimum weekly FIT batch analysis would minimise the potential of this degradation to misclassify results.

Our serial sample data provides some insights into sample variability and enables some comment on sample stability. It is well established that Hb degrades in faecal material (10) and this presents a potential for false negatives as the concentration may fall beneath that considered positive or detected. In the serial samples from patients we do not know the magnitude of degradation in individual specimens but we found no statistical difference with respect to categorical positive or negative results with this time series of specimens. Our analysis is limited in that the intervals between specimens are variable. It is possible that the variability between days (a component of biological variability) and from sampling (a component of analytical variability) may be larger than the concentration changes associated with degradation: if sample degradation were the dominant variable it would be expected that the time series analysis would be significant.

### Comparisons to previous literature

In a previous study (15) of the diagnostic accuracy of one vs two specimens it was concluded that two specimens offered no advantage over one specimen. Sequential results of 400 µg/g were sometimes followed by <10 µg/g from the same patient taken a median of 6 days apart. It is not certain whether these differences are due to biological variation, inconsistent bleeding or sampling imprecision. A study (16) of the utility of two sample strategies for colorectal cancer in a symptomatic population it was noted that there was 39.2% non-concordance between the first FIT result and the maximum FIT result. It would therefore appear that whatever strategy is used, bleeding from a lesion may be inconsistent and any single sample will, on average, detect with the same degree of diagnostic accuracy. Of note, previous studies of FIT in symptomatic patients have noted a small number of false negatives (4). In a recent study (17) using FIT in a symptomatic population in a 2 week wait pathway 12.5% of colorectal cancers were missed using a single patient collected specimen using a threshold of 10 µg/g. The method utilised was different from the one utilised in our laboratory study, the OC-SensorTM (Eiken Chemical Company, Tokyo, Japan) and had a reported detection limit of 4 µg/g. As with other studies it is not certain whether these false negative results were due to variable patterns of bleeding, poor sampling by the patient, or another cause. The study also noted that the majority of FIT results from patients with colorectal cancer had very high values (>150 µg/g) which would suggest these cases should be referred with higher priority.

### Implications for research and practice

There is on-going debate as to how FIT results should be reported from those that consider the result as “detected”/”not detected” through to quantitative reporting of numeric values (4,7). Given the high imprecision of analysis when sampling is taken into consideration we would caution against quantitative reporting of faecal Hb due to the high uncertainty of measurement. Further audit of larger datasets derived from symptomatic patients in routine clinical practice are required to provide evidence of the association between Hb concentration and clinical outcomes and to further inform qualitative reporting strategies. The binary nature of reporting that we suggest may make for more straightforward pathways for the triage of patients who have been deferred due to the COVID-19 pandemic and require re-prioritisation into urgent and less urgent groups.

The quality assurance of faecal tests, including FIT, presents unique analytical challenges compared to most clinical specimens used for diagnostic testing, such as whole blood, serum, plasma or urine. This is demonstrable in the lack of verification data in native faecal material available from manufacturers supplying sampling devices. Existing data, depend upon synthetic material. It also presents challenges to laboratories with respect to both IQC and EQA. In our laboratory, we have commenced analysis of samples collected directly by patients into the collection device and noted that, despite collection guidance leaflets, a YouTube video and targeted training of staff advising patients, many samples arrive with faecal material clearly on the outside of the container and no specimen apparent in the buffer. Anecdotally, we have noted in many of the samples where sample integrity is questionable from visual inspection of the collection device, the patient is over 75 years of age.

Collection of samples in patients where there has been a change in bowel habit represents specific additional challenges and stool water content can vary between 60 and 82% (18). The dilutional effect of this will impact on quantitative measures and the lack of form will affect entrapment of the specimen itself in the manufacturer’s collection device. We have noted a minority of specimens where considerably more than a 30 second vortex is required to suspend the faecal matter into the buffer. Protocols for how to manage these challenging specimens observed in the symptomatic population are lacking. Furthermore there is considerable concern about risk of faecal-oral or aerosol transmission of COVID-19 (19) and the risks of sample manipulation and the need for a period of very high throughput need to be balanced against analytic accuracy.

The evaluation of collection device robustness is particularly important to continue. One detailed study has reported that collection devices can be used effectively even when not used as recommended by the manufacturers (20). Whilst it demonstrated significant differences in the absolute FIT concentration the effect on diagnostic performance was small. As with our study the testing involved specimen homogenisation and sampling in a laboratory to reduce variability.

## Concluding remarks

In summary we have found that a commonly used FIT method shows good consistency of categorising Hb results against the DG30 threshold, and therefore appears suitable for this clinical application. However, there appear numerous sources of variability that require further investigation and optimisation. It is therefore important that all FIT studies identify the relative contributions of biological variation, sampling technique, Hb stability and method performance to false negative results as these are consistently observed in the literature. The high imprecision precludes reporting of quantitative FIT concentrations and on current analytical performance we consider a categorical approach, for example negative or positive relative to recommendation of a 2 week wait referral is more appropriate, potentially simplifying pathways to reprioritise patients with lower GI cancer symptoms who have had definitive diagnosis delayed by the COVID-19 pandemic.

## Data Availability

All data is available through Dr Brian Shine.

## Acknowledgements

James E. East was funded by the National Institute for Health Research (NIHR) Oxford Biomedical Research Centre. Brian D Nicholson is an NIHR Academic Clinical Lecturer and is supported by the NIHR Oxford Medtech and In-Vitro Diagnostics Co-operative. The views expressed are those of the author(s) and not necessarily those of the National Health Service, the NIHR or the Department of Health.

## References

1. World Health Organization. Colorectal cancer Source: Globocan 2018 Number of new cases in 2018, both sexes, all ages [Internet]. 2018. [cited 30 May 2019]. Available: http://gco.iarc.fr/today/data/factsheets/cancers/10_8_9-Colorectum-fact-sheet.pdf

2. Wilkins T, McMechan D, Talukder A. Colorectal Cancer Screening and Prevention. Am Fam Physician. 2018 May 15;97(10):658–665.

3. National Institute for Health and Care Excellence. Quantitative faecal immunochemical tests to guide referral for colorectal cancer in primary car. Diagnostics guidance (DG30). 2017. https://www.nice.org.uk/guidance/dg30 (Accessed 8 Mar 2020).

4. Mole G, Withington J, Logan R. From FOBt to FIT: making it work for patients and populations. Clinical Medicine 2019 Vol 19, No 3: 196–9

5. Nicholson BD, James T, East JE et al. Experience of adopting faecal immunochemical testing to meet the NICE colorectal cancer referral criteria for low-risk symptomatic primary care patients in Oxfordshire, UK. Frontline Gastroenterol 2018

6. National Institute for Health and Care Excellence. Suspected cancer: Recognition and referral. NICE guideline (NG12, 2017. [updated 2017]. https://www.nice.org.uk/guidance/ng12. (Accessed 8 Mar 2020).

7. Fraser CG, Benton SC. Detection capability of quantitative faecal immunochemical tests for haemoglobin (FIT) and reporting of low faecal haemoglobin concentrations. Clin Chem Lab Med. 2019;57:611–616.

8. Godber IM, Benton SC, Fraser CG. Setting up a service for a faecal immunochemical test for haemoglobin (FIT): a review of considerations, challenges and constraints. J Clin Pathol. 2018;71:1041–1045.

9. Carrol M, Piggott C, Pearson S et al Evaluation of quantitative faecal immunochemical tests for haemoglobin. Dec 2014. http://194.97.148.137/assets/downloads/pdf/activities/fit_reports/gmec_fit_evaluation_report.pdf

10. Brown LF, Fraser CG. Effect of delay in sampling on haemoglobin determined by faecal immunochemical tests. Ann Clin Biochem. 2008;45:604–5.

11. Steele R, Forgacs I, McCreanor G, Benton S, Machesney M, Rees C, et al. Use of faecal occult blood tests in symptomatic patients. BMJ. 2015;351

12. Hamilton W, Hajioff S, Graham J, Schmidt-Hansen M. Authors’ reply to Steele and colleagues. BMJ: British Medical Journal. 2015;351.

13. Nicholson BD, Oke J, Friedemann Smith C, Phillips JA, Lee J, Abel L, Kelly S, Gould I, Mackay T, Kaveney Z, Anthony S, Hayles S, Lasserson D, Gleeson F. The Suspected CANcer (SCAN) pathway: protocol for evaluating a new standard of care for patients with non-specific symptoms of cancer. BMJ Open. 2018;8(1):e018168. doi: 10.1136/bmjopen-2017-018168.

14. Gratton J, Phetcharaburanin J, Mullish BH, Williams HR, Thursz M, Nicholson JK, Holmes E, Marchesi JR, Li JV. Optimized Sample Handling Strategy for Metabolic Profiling of Human Feces. Anal Chem. 2016; 88: 4661–8.

15. Turvill J, Mellen S, Jeffery L, Bevan S, Keding A, Turnock D. Diagnostic accuracy of one or two faecal haemoglobin and calprotectin measurements in patients with suspected colorectal cancer. Scand J Gastroenterol. 2018;53:1526–1534.

16. Auge JM, Fraser CG, Rodriguez C, Roset A, Lopez-Ceron M, Grau J, Castells A, Jimenez W. Clinical utility of one versus two faecal immunochemical test samples in the detection of advanced colorectal neoplasia in symptomatic patients. Clin Chem Lab Med. 2016; 54:125–32.

17. Chapman C, Bunce J, Oliver S, Ng O, Tangri A, Rogers R, Logan RF, Humes DJ, Banerjea A. Service evaluation of faecal immunochemical testing and anaemia for risk stratification in the 2-week-wait pathway for colorectal cancer. BJS Open. 2019;3:395–402.

18. Blake MR, Raker JM, Whelan K. Validity and reliability of the Bristol Stool Form Scale in healthy adults and patients with diarrhoea-predominant irritable bowel syndrome. Aliment Pharmacol Ther. 2016; 44:693–703.

19. Cheung KS, Hung IF, Chan PP, Lung KC, Tso E, Liu R, Ng YY, Chu MY, Chung TW, Tam AR, Yip CC, Leung KH, Yim-Fong Fung A, Zhang RR, Lin Y, Cheng HM, Zhang AJ, To KK, Chan KH, Yuen KY, Leung WK. Gastrointestinal Manifestations of SARS-CoV-2 Infection and Virus Load in Fecal Samples from the Hong Kong Cohort and Systematic Review and Meta-analysis. Gastroenterology. 2020 Apr 3. pii: S0016-5085(20)30448-0. doi: 10.1053/j.gastro.2020.03.065. [Epub ahead of print]

20. Gies A, Gruner LF, Schrotz-King P, Brenner H. Effect of imperfect compliance with instructions for fecal sample collection on diagnostic performance of 9 fecal immunochemical tests. Clin Gastroenterol Hepatol. 2019;17:1829–1839.

